# Screening for SARS-CoV-2 infections with colorimetric RT-LAMP and LAMP sequencing

**DOI:** 10.1101/2020.05.05.20092288

**Authors:** Viet Loan Dao Thi, Konrad Herbst, Kathleen Boerner, Matthias Meurer, Lukas PM Kremer, Daniel Kirrmaier, Andrew Freistaedter, Dimitrios Papagiannidis, Carla Galmozzi, Steffen Klein, Petr Chlanda, Dina Khalid, Isabel Barreto Miranda, Paul Schnitzler, Hans-Georg Kräusslich, Michael Knop, Simon Anders

## Abstract

The COVID-19 pandemic caused by the novel SARS-CoV-2 virus poses a significant public-health problem. In order to control the pandemic, rapid tests for detecting existing infections and assessing virus spread are critical.

Approaches to detect viral RNA based on reverse transcription loop-mediated isothermal amplification (RT-LAMP) hold outstanding promise towards greatly simplified and broadly applicable testing methods. RT-LAMP assays appear more robust than qPCR-based methods and only require incubation at a constant temperature, thus eliminating the need for sophisticated instrumentation.

Here, we tested a two-color RT-LAMP protocol using clinical SARS-CoV-2 samples and also established a protocol that does not require prior RNA isolation (“swab-to-RT-LAMP”). Our study is based on several hundred clinical patient samples with a wide range of viral loads, thus allowing, for the first time, to accurately determine the sensitivity and specificity of the RT-LAMP assay for the detection of SARS-CoV-2 in patients. We found that RT-LAMP can reliably detect SARS-CoV-2 samples with a qPCR threshold cycle number (CT value) of up to 30 in the standard RT-qPCR assay. We used both, either purified RNA or direct pharyngeal swab specimens and showed that RT-LAMP assays have, despite a decreased sensitivity compared to RT-qPCR, excellent specificity. We also developed a multiplexed LAMP-sequencing protocol as a diagnostic and validation procedure to detect and record the outcome of RT-LAMP assays. LAMP-sequencing is fully scalable and can assess the results of thousands of LAMP reactions in parallel. Finally, we propose applications of RT-LAMP based assays for SARS-CoV-2 detection.

## Introduction

The outbreak of a pandemic caused by a new coronavirus, SARS-CoV-2 (Zhu et al., 2020), poses major challenges for national health care systems. A yet rather unspecified proportion of people, especially the elderly and those with pre-existing conditions, are at high risk of a severe course of the SARS-CoV-2-induced disease COVID-19 (Phua et al., 2020). This can lead to a high burden on the health care system resulting in a situation where not all patients can receive adequate treatment. Further, due to limited capacities, only people with symptoms are usually being tested, although recent studies confirmed that many SARS-CoV-2 carriers are asymptomatic (Mizumoto et al., 2020, Arons et al., 2020). This suggests that infection-control strategies focussing on symptomatic patients are not sufficient to prevent virus spread.

Therefore, large scale diagnostic procedures are needed to determine the spread of the virus in the population quickly, comprehensively, and sensitively. This allows for the rapid isolation of infected persons during an existing wave of infection and helps to identify newly emerging outbreaks; and further for measures to slow down the pandemic, to adapt the capacity of the health care system to the expected number of patients, and distribute essential resources. Moreover, continuous and repeated testing for acute infections of large groups within a population will be required as a long-term perspective to contain new outbreaks while keeping societies and economies functional until efficient vaccines become available.

An active SARS-CoV-2 infection can be diagnosed by detecting the viral genome or viral antigens in patient samples. An assay for the latter is limited by the sensitivity, specificity, and production speed of diagnostic antibodies, while detecting viral genomes only requires specific oligonucleotides, which facilitates testing of large cohorts.

The SARS-CoV-2 diagnostic pipeline which has proven to be successful and which is currently used in many test centers consists of three steps: collecting nasopharyngeal or oropharyngeal swab specimens, isolation of total RNA, and specific detection of the viral genome by RT-qPCR which includes a reverse transcriptase (RT) step, translating the viral RNA into DNA, followed by a semi-quantitative DNA polymerase chain reaction (qPCR) using oligonucleotides specific for the viral cDNA (RT-qPCR). As a result, a short piece of the viral genome is strongly amplified, which is detected by a sequence-specific oligonucleotide-probe labelled with a fluorescent dye.

This procedure includes several steps that require liquid handling; therefore, the detection process in a clinical diagnostics laboratory takes about 3 to 24 hours or more, depending on the sample volume and process optimization of the test center. In addition, in the context of the pandemic, many of the reagents required are only slowly being replenished due to insufficient production capacity or lack of international transport. Therefore, increasing daily test capacities based on RT-qPCR-based SARS-CoV-2 diagnostics is limited. In order to accelerate and optimize diagnostics, new, scalable methods for RNA isolation and the detection of viral genomes are needed.

An alternative to RT-qPCR is reverse transcription loop mediated isothermal amplification (RT-LAMP) (Notomi et al., 2000; Nagamine et al., 2002; Tomita et al., 2008). RT-LAMP reactions include a reverse transcriptase as well as a DNA polymerase with strong strand displacement activity and tolerance for elevated temperatures, and up to six DNA oligonucleotides of a certain architecture. Samples with potential template molecules are added to the reaction and incubated for 20-60 minutes at a constant temperature (e.g. 65°C). The oligos act as primers for the reverse transcriptase and additional oligos for the DNA polymerase that are designed so the DNA products loop back at their ends. These in turn serve as self-priming templates for the DNA polymerase. In the presence of a few RNA template molecules, a chain reaction is set in motion, which then runs until the added reagents (e.g., dNTPs) are used up.

In order to detect the DNA production in LAMP assays, various approaches have been described in the literature. One possibility is to use a pH indicator (e.g. phenol red) and run the reaction in a weakly buffered environment. As the chain reaction proceeds, the pH lowered, which results in a visible color change from red to yellow making it an especially appealing assay for point-of-care diagnostics (Tanner et al., 2015). Previously, RT-LAMP assays have been proposed for diagnostic detection of other RNA viruses, such as e.g. influenza (Ito et al., 2006). Also, several preprints demonstrate the use of isothermal DNA amplification to detect small amounts of SARS-CoV-2 RNA. The majority of these studies used *in vitro* transcribed short fragments of the viral genomic RNA (Yu et al., 2020; Zhang et al., 2020; El-Tholoth et al., 2020) and showed a detection limit of somewhere between 10-100 RNA molecules per reaction.

We therefore set out and assessed the sensitivity and selectivity of the RT-LAMP assay to detect SARS-CoV-2 infections from clinical RNA preparations as well as directly from pharyngeal swab specimens (termed “swab-to-RT-LAMP assay”) from COVID-19 patients. Here, we used a sufficient sample size with a wide viral load range allowing for the first time to determine accurately the sensitivity range of the colorimetric RT-LAMP assay for the detection of SARS-CoV-2 in patients. An overview of the samples available to us is shown in Figure 1.

**Figure 1.**
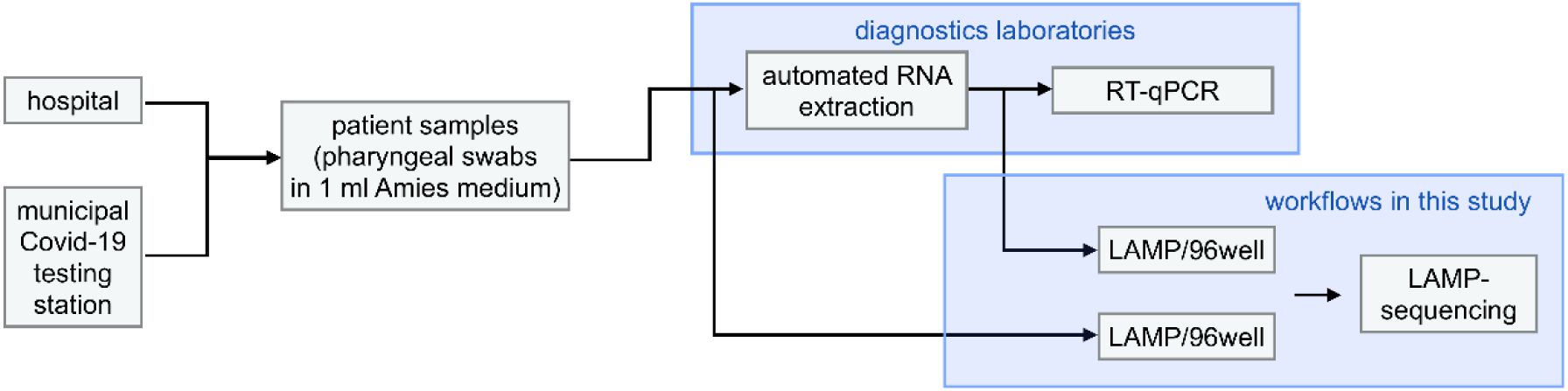
SARS-CoV-2/COVID-19 samples used in this study. Samples available for this study were derived from COVID-19 tests from the hospital or a municipal testing station and surplus material was passed on to us, usually within a few hours to one day after the hospital diagnostic laboratory had removed an aliquot for qPCR analysis. Aliquots of the samples were used for individual assays or for seed plate generation (96-well format) for multiplexed analyses.

The performance of an RT-LAMP test does not require expensive special equipment such as a thermal cycler with real time fluorescence measurement, as positive samples are determined by a color change in the sample from red to yellow within 30 min after start of the incubation at 65°C. For detection, simple mobile phone cameras, copy machines, office scanners to spectrophotometric quantification can be used. We then describe a multiplexed sequencing protocol using barcoded tagmentation that enables rapid identification of correct and false positive reactions in many thousands of RT-LAMP assays within the same next-generation sequencing run, e.g. using Illumina dye or Oxford nanopore sequencing.

Together, the results of this study allow a critical evaluation of the RT-LAMP colorimetric reaction for the detection of SARS-CoV-2 RNA in patient samples. Our data show the limitations of the RT-LAMP method, but also the possibilities and different applications of a simplified hot swab-to-RT-LAMP’ procedure, which does not require any purification of the swab specimen

## Results

### Sensitivity of colorimetric RT-LAMP assay using artificial SARS-CoV-2 RNA template

To validate the detection of SARS-CoV-2 RNA RT-LAMP, we used the WarmStart Colorimetric RT-LAMP 2X Master Mix (DNA & RNA) from New England Biolabs. Several primer sets were recently proposed for RT-LAMP-based detection of SARS-CoV-2 by Zhang et al., (2020) as well as by Yu et al., (2020) and subsequently validated with *in vitro* translated RNA. We prepared and tested two primer sets for different RNA sections of the SARS CoV-2 genome, namely Zhang et al’s set N-A (targeting N gene) and their set 1a-A (targeting ORF 1a gene) (Table 1, Materials and Methods). Figure 2 shows that the oligo set for the N gene is capable of detecting 100 *in vitro* transcribed (IVT) RNA molecules in a test reaction with 1 microliter (µl) of RNA solution. To put this into perspective: Wölfel et al., (2020) found that COVID-19 patients showed up to 7×10^8^ RNA copies per throat swab in the first week after infection. Assuming a quantitative extraction of all RNA molecules in the sample, this should equal a concentration of up to several million RNA molecules per 1 µl of RNA extract, depending on the efficiency and concentration factor of the used RNA extraction protocol. Thus, the RT-LAMP reaction should be sufficiently sensitive for a reliable detection of SARS CoV-2 viruses even in patients with a much lower pharyngeal viral load.

**Figure 2:**
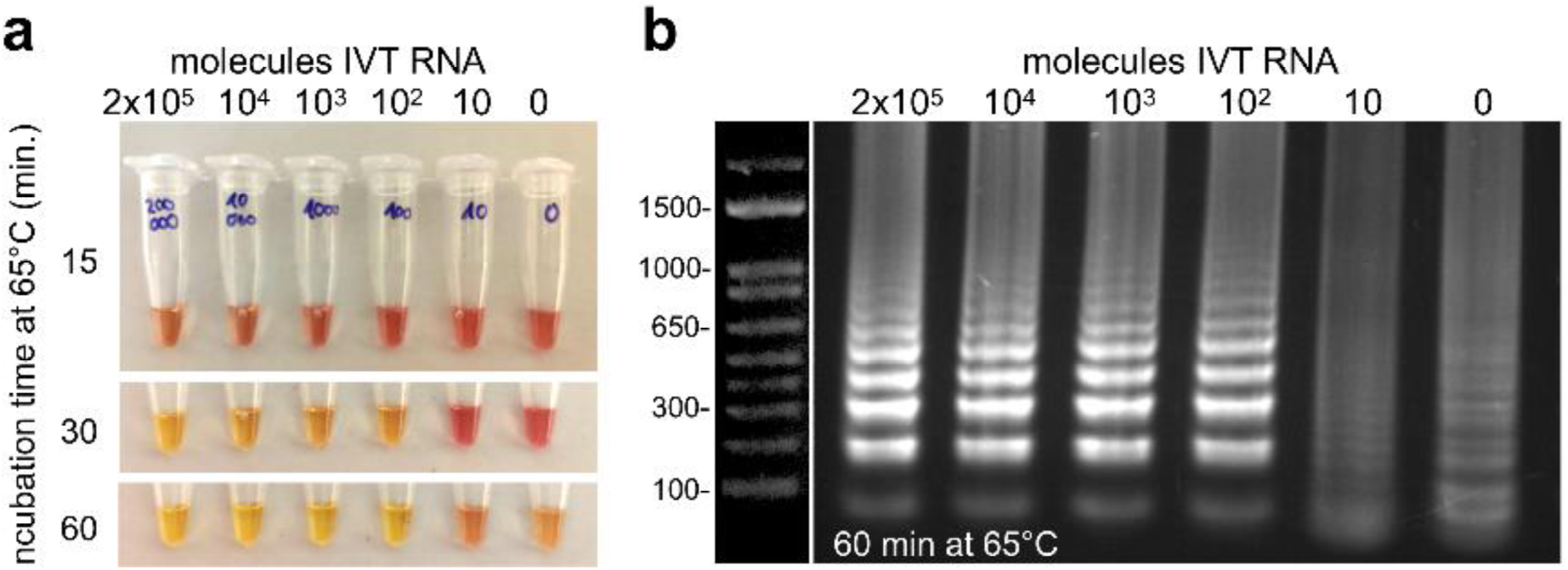
Sensitivity of the RT-LAMP test with IVT RNA. (**a**) Defined numbers of *in vitro* transcribed (IVT) RNA molecules of N gene were added to the reaction and incubated at 65°C. At indicated times, samples were taken out of the heating block, cooled on ice to stop the reaction, and photographed. (**b**) 5 µl of the RT-LAMP reaction product (after 60 min) were analysed on a 2% agarose gel. The typical band pattern of a successful RT-LAMP reaction was visible in the samples with 100 or more RNA molecules, i.e., in those samples that showed a color change after 30 minutes. Upon prolonged incubation at 65°C, in the case of the colorimetric assay for timepoints > 30-35 min, the negative control becomes yellowish and non-specific DNA smear appears when loaded on a gel. This is a well known problem with RT-LAMP (Gadkar et al., 2018).

To further establish the sensitivity and stability of RT-LAMP reagents and assay, we also explored storage of the premixed reaction at different temperatures, temperature optima of the RT-LAMP assay, and sensitivity of different primer sets (see Methods and Supplementary Discussion and Comments). In conclusion, the primer set N-A for the N gene appears to provide the best sensitivity for detection of RNA. Premixed primer/RT-LAMP reagents can be stored at −20° overnight (but not longer), and they should not be stored for more than 30 to 60 min on ice before use. We also noticed that exposure to air (open tubes) leads to an acidification of the reaction mix, which, due to its low buffer capacity, rapidly changes its color.

### Establishing colorimetric RT-LAMP test with clinical samples

For evaluation of the RT-LAMP test procedure with patient samples, its sensitivity and specificity has to be compared with a validated RT-qPCR method.

To do so, we first compared the detection of viral RNA in 95 human RNA extracts from pharyngeal swabs both by RT-qPCR and by the RT-LAMP assay using primer sets against ORF 1a and the N gene (Figure 3). To visualize the RT-LAMP reaction, the photo-function of a normal mobile phone is sufficient (Figure 3a). To quantify the reaction, the difference in absorbance (ΔOD) of the samples at 434 nm and 560 nm (corresponding to the absorbance maxima of the two forms of phenol red that is used in the RT-LAMP assay as pH-sensitive dye) was measured. For measurement a Tecan Spark Cyto plate scanner was used, but any plate scanner that allows OD measurements at different wavelengths can be used. ΔOD values were compared in a scatter plot with the CT values of the standard RT-qPCR of the hospital laboratory’s routine diagnostic pipeline (which used primers against the E gene) (Figure 3b). The used RT-LAMP enzyme mix contains oligonucleotide-based aptamers that function as reversible inhibitors for the RTx (reverse transcription xenopolymerase) and Bst 2.0 (strand-displacing polymerase) enzyme present in the reaction. This allowed stopping of the reaction at different timepoints by cooling the samples on ice and inspecting the color visually (Figure 3a) or by measuring ΔOD at a given time point of incubation at 65°C (e.g. ΔOD_30min_) (Figure 3c). We noted that interruption of the reaction by cooling on ice for extended periods of up to 20 min is possible without loss of sensitivity. Usually the color change is detected in positive samples at timepoints > 10 min, and all positive samples have changed at 30 min. Further incubation led to color changes in negative samples. Inspection of the color curves revealed a wide gap between positive and negative samples, indicating that measurement in the time window from 25 to 30 min yields very robust results: negative samples had an absorbance difference below 0 and positive an absorbance difference well above +0.3 (Figure 3b).

**Figure 3:**
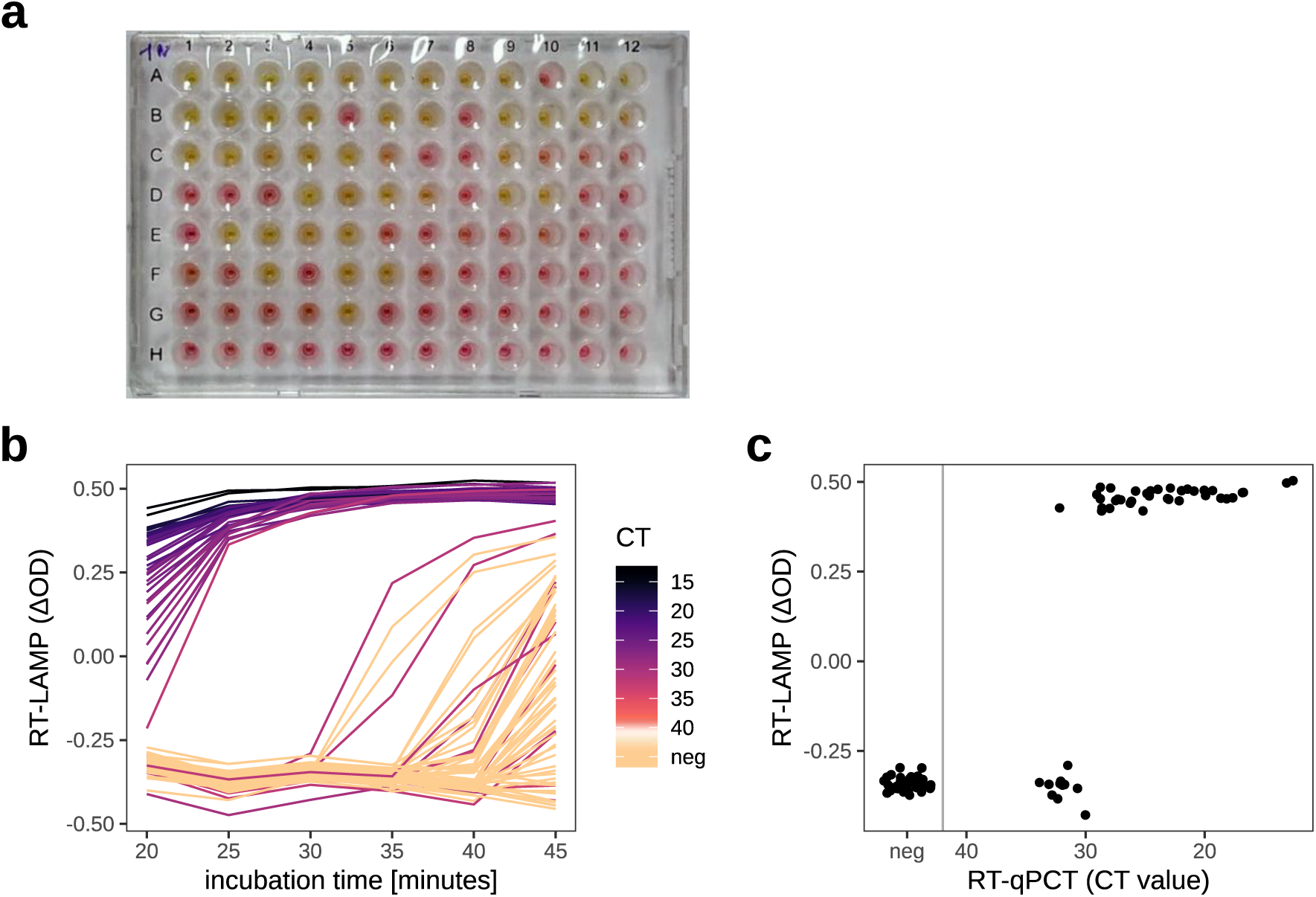
Sensitivity and specificity of the colorimetric RT-LAMP test with clinical samples in comparison to RT-qPCR. (**a**) Isolated RNA from 95 pharyngeal swabs (Figure 1) were processed for a RT-LAMP reaction. The RT-LAMP reaction was incubated at 65°C, stopped at different time points by cooling on ice for 30 sec, and the plate was photographed with a mobile phone. The image shown was taken 30 min after the start of the incubation at 65 C. Wells with yellow color: color change due to successful RT-LAMP amplification of a fragment of SARS-CoV-2 N gene (oligos N-A, see Methods), (**b**) Quantification of the color change in all wells using spectrophotometric OD measurements of the color change at two wavelengths 434 nm and 560 nm at the given time points. The color change is given as the difference between the absorbances at the two wavelengths (ΔOD = OD_434 nm_ - OD_560 nm_). Yellow, positive samples yield a ΔOD of approximately 0.3-0.4. Each line represents one sample, the line color codes for the sample’s CT value from RT-qPCR (see legend) (c) Comparison of the RT-LAMP results from (b) for time point 30 minutes with the CT values from RT-qPCR.

Comparison of RT-LAMP positive samples with the CT values from RT-qPCR showed that for N-A oligos all patient samples with a CT < 30 showed a color change in the RT-LAMP test, whereas for CT values between 30-35, a color change was observed for 1/10 patient samples. We also tested other oligos, e.g. oligos against ORF1a gene, which revealed a lower sensitivity of detection (Supplementary Figure 1). We concluded that the RT-LAMP test with the primer set against the N gene could robustly detect SARS-CoV-2 infection in patient samples with a CT value below 30 and we decided to use this primer set for all further experiments.

### Validation of colorimetric RT-LAMP for SARS-CoV-2 testing using a large cohort

In order to ensure the specificity and selectivity of the test, it must be demonstrated that the test works with RNA samples obtained on different RNA extraction platforms, and that it is robust against the subject-to-subject heterogeneity of material obtained from pharyngeal swabs. Therefore, we analyzed further RNA samples, to a total of 792 pharyngeal swabs taken from the hospital laboratory’s clinical routine tests, for which RNA had been extracted using different platforms (QiaSymphony and QiaCube, Qiagen). A visualisation of the data, again taken 30 minutes after the start of the incubation at 65°C, showed comparable behaviour of the samples on all ten plates that were run (represented by different colors in Figure 4a). This meant that none of the extraction protocols affected the RT-LAMP test significantly.

**Figure 4:**
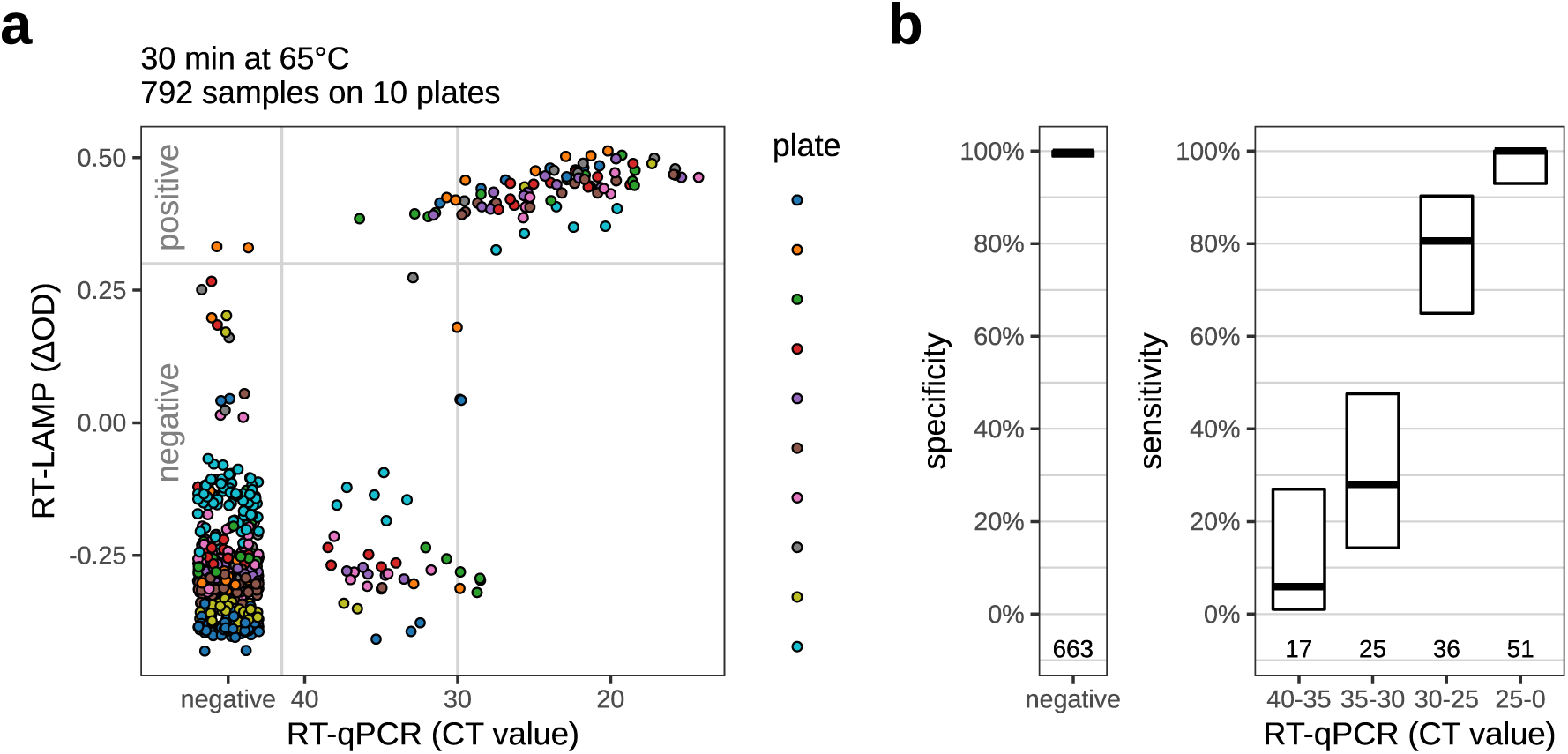
Detection of SARS-CoV-2 RNA in patient samples using the RT-LAMP assay. (**a**) Scatter plot to visualize all samples across the ten test plates. RT-LAMP quantification as shown in Figure 3c. Time point of measurement: 30 min after the start of the 65°C incubation. See Supplementary Table S1 for counts. The plate shown in Figure 3 is not included here. (**b**) Sensitivity and specificity of the RT-LAMP assay, as inferred from the data shown in (a). Left: The specificity (i.e., the fraction of qPCR negatives correctly identified as negative by the RT-LAMP assay) and its 95% CI. Right: Blackhorizontal lines give, for several ranges of CT values, the sensitivity of the test (i.e., the proportion of samples within this CT range that the test detects as positive). The gray boxes indicate the 95% confidence intervals (Wilson’s binomial CI) for these values. Sample numbers are indicated at the bottom.

We considered an absorbance difference above +0.3 as a positive and equal or below +0.3 as a negative result. Overall, RT-qPCR positive samples with a CT-value < 30 scored positive in RT-LAMP assay, whereas samples with CT values between 30 and 40 mostly scored negative. We observed small differences between different plates on the exact sensitivity threshold, probably caused by slight variability in plate or reagent handling or individual samples. We observed two RT-qPCR negative samples that scored positive in the RT-LAMP (Figure 4a). The overall specificity of the test was 99.7% (Wilson’s 95% confidence interval: 98.9% - 99.9%), the sensitivity for samples with CT < 30 was 92.0% (Wilson’s 95% confidence interval: 84.3% - 96.0%). For details, see Figure 4b and Supplementary Table S1.

### Multiplexed sequencing of RT-LAMP products (LAMP-sequencing)

So far the results indicated that the colorimetric RT-LAMP offers a robust readout after 25–30 min incubation at 65°C. However, some negative samples occasionally also turned yellow upon prolonged incubation (Figure 2a). It is known that RT-LAMP reactions can occasionally generate spurious amplicons, especially in the absence of target sequences and upon prolonged incubation, resulting in false positive results (Gadkar et al., 2018). By electrophoresis, these samples do not show the characteristic ladder for a successful RT-LAMP (Figure 2b).

A scalable approach to further verify RT-LAMP results unambiguously would hence be useful. LAMP reactions yield DNA products of heterogeneous structure which is why we needed to use a sequencing strategy that is as unbiased as possible as well as fully scalable in order to allow the analysis of the large number of samples involved. To this end, we established LAMP-sequencing, a fully scalable multiplexed sequencing strategy using Tn5 tagmentation. This allowed us to enrich LAMP-specific amplicons without selecting for a certain subset of LAMP products. We used a set of 96 barcoded adapters to barcode the RT-LAMP reaction products in 96-well plates. These adapters also contained unique molecular identifiers (UMIs) for quantification purposes (Kiviola et al., 2011). After tagmentation, the barcoded fragments were pooled and size-selected by bead purification to remove excess adapters. A second set of barcoded primers was then used to amplify the pooled tagmented RT-LAMP fragments (Figure 5a). Analysis of the sequences allowed us to unambiguously identify for each well whether the RT-LAMP reaction yielded a product that incorporated the viral genomic sequence.

**Figure 5:**
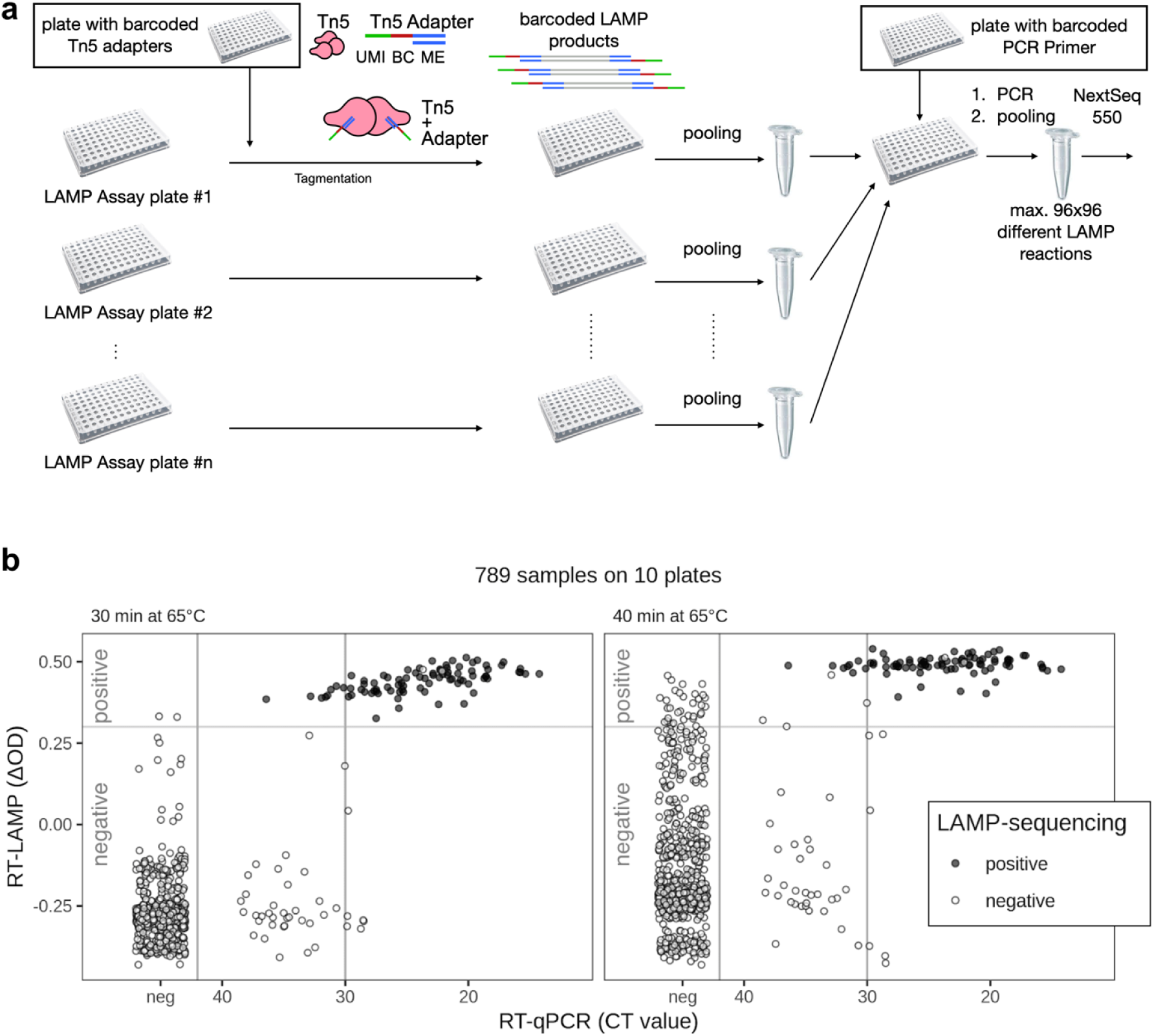
Multiplexed sequencing of RT-LAMP products (LAMP-sequencing) (**a**) Workflow for LAMP-sequencing. We used a plate of 96 barcoded adaptors that included unique molecular identifiers (UMIs) for Tn5 tagmentation of the RT-LAMP products for each 96-well RT-LAMP reaction. Tagmentation involves one pipetting step per Rt-LAMP reaction plate, before the samples from each plate can be pooled. After purification and size selection of tagmented fragments, one PCR per plate is used to introduce a second barcode, one for each plate. Afterwards, the samples were analyzed using an Illumina NextSeq 550 run. (**b**) Comparison of RT-LAMP, RT-qPCR, and sequencing results for all samples detected during sequencing. The left panel shows the results for a total of ten plates, comprising the same plates as shown in Figure 4. The sequencing outcome is color-coded: black if a substantial number of reads contained SARS-CoV-2 sequence, white otherwise. While the RT-LAMP test was scored after 30 minutes of incubation, the sequencing was performed after the samples had been incubated for 40 minutes (which yields the spurious LAMP reactions in negative samples, as shown by the high ΔOD value for many of them). Therefore, we show the RT-LAMP score after both 30 minutes (left) and 40 minutes (right) total incubation time. For statistical parameters, see Supplementary Table S2.

LAMP-sequencing yielded results for 789 of the 792 samples previously evaluated by RT-LAMP, confirming almost all of the findings of the colorimetric scoring of the RT-LAMP assay (Figure 5, Supplementary Figure S2, Table 2). All samples which appeared positive in LAMP-sequencing also scored positive in the RT-LAMP and RT-qPCR assays, indicating an exceptional precision. LAMP-sequencing is directly dependent on the RT-LAMP reaction. We observed two negative samples in the sequencing which had scored positive in the RT-LAMP reaction, indicating that these were false positives of the colorimetric RT-LAMP assay (compare Figure 4a and Figure 5a). Overall, we could show that LAMP-sequencing can serve as a powerful validation tool for a large number of RT-LAMP assays in high-throughput. In addition, LAMP-sequencing confirmed our high threshold for positive RT-LAMP samples as spurious amplification in some RT-LAMP samples did result in a weak colorimetric shift but not in detectable virus sequences.

### Swab-to-RT-LAMP assays using crude specimen

RNA isolation is time-consuming, costly, and depends on reagents with potentially limited supply in the course of a pandemic. Alternative, non-commercial solutions for RNA isolation, e.g. using silica gel matrix or magnetic beads, require specialized knowledge and cannot be implemented easily for point-of-care or decentralized patient screening.

Several reports indicate that RT-qPCR (Bruce et al., 2020, Smyrlaki et al., 2020, Wee et al., 2020) and RT-LAMP assays (Lamb et al., 2020, Rabe and Cepko 2020) are compatible with direct testing of nasopharyngeal and oropharyngeal swab specimens without a prior purification or extraction step (in the following termed ‘swab-to-RT-LAMP’ for RT-LAMP assays), but these studies relied on detecting spiked-in IVT RNA. Towards establishing RT-LAMP assays with crude specimens, we first wanted to assess the stability of ‘naked’ RNA in swab specimens, which were collected in Amies medium (eSwab, Copan Italia). To this end we titrated defined numbers of IVT RNA molecules of SARS-CoV-2 N gene into swab samples from COVID-19 negative control subjects. We tested different conditions, in particular the influence of detergent (to inactivate the virus) and heat (to denature the capsid and release the viral RNA as well as inactivate the virus). Heat treatment has been LAMP assays with crude specimens, we first wanted to assess the stability of ‘naked’ RNA in swab specimens, which were collected in Amies medium (eSwab, Copan Italia). To this end we titrated defined numbers of IVT RNA molecules of SARS-CoV-2 N gene into swab samples from COVID-19 negative control subjects. We tested different conditions, in particular the influence of detergent (to inactivate the virus) and heat (to denature the capsid and release the viral RNA as well as inactivate the virus). Heat treatment has been shown to be beneficial for testing swab specimens without RNA isolation in RT-qPCR assays (Fomsgaard et al., 2020).

As shown in Figure 6a, IVT RNA when spiked into swabs taken from control (COVID-19 negative) test subjects, is hardly detected by RT-LAMP, most likely indicating degradation of the RNA. Neither the addition of SDS or a heat-stable RNAse inhibitor prevented this rapid degradation. Similarly, IVT RNA added before heat incubation of swab specimens at 95°C for 5 mins failed to be detected by a RT-LAMP assay, with slight differences depending on the swab specimen tested (Figure 6b). However, when DNA instead of RNA was used, much higher stability was observed (Figure 6c).

**Figure 6:**
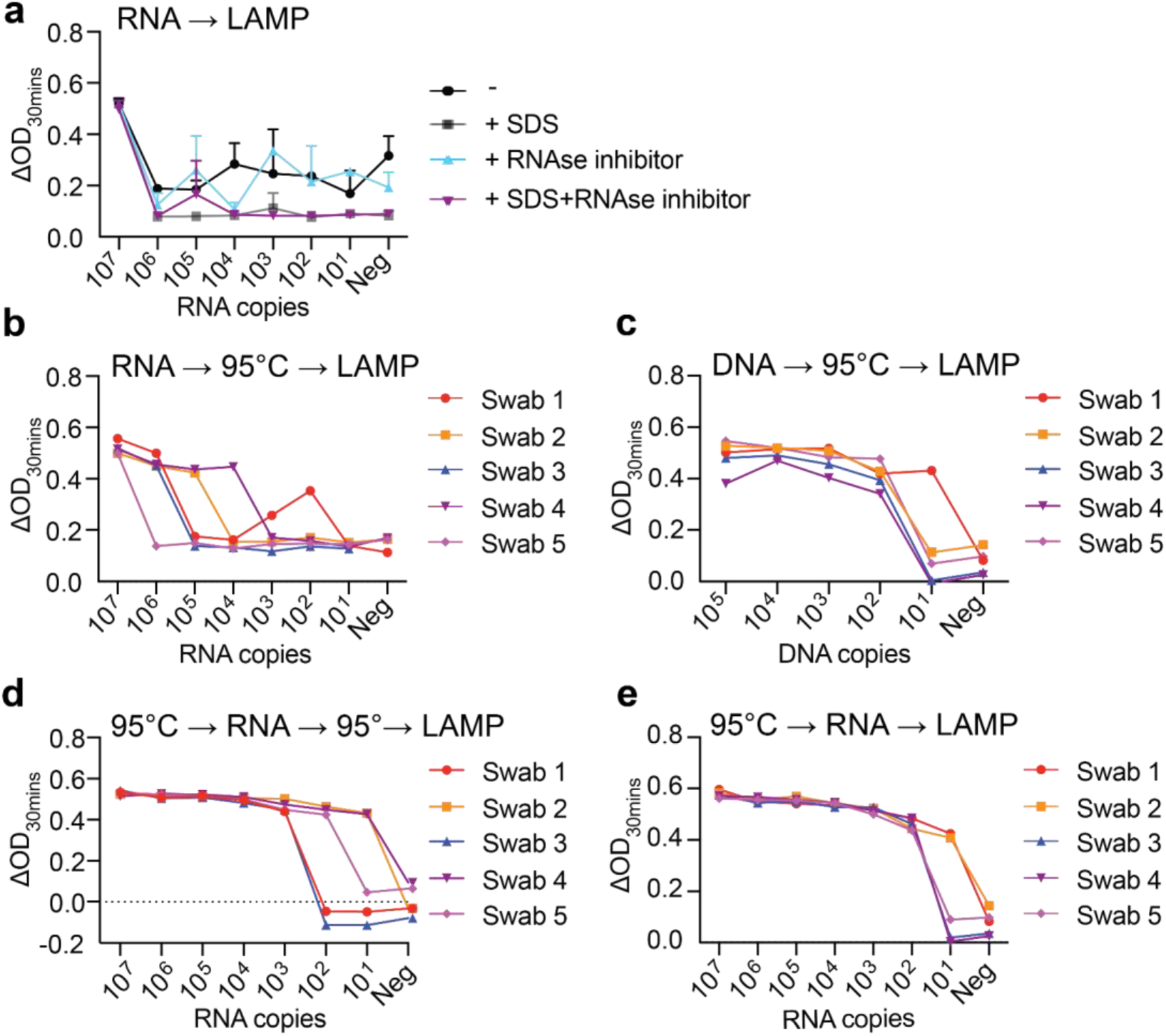
RNA stability and limit of detection of RT-LAMP assay in pharyngeal swab specimens with IVT RNA. SARS-CoV-2 RNA titration in control specimen by RT-LaMP (**a**) Defined numbers of molecules of IVT RNA of gene N were added to aliquots of control specimen adjusted or not to 0.1% SDS and/or treated with RNAse inhibitor as indicated, n=2. Control specimens from different negative patients were spiked with RNA or DNA before (**b** and **c**), between (**d**) or after (**e**) heat treatment (95°C / 5 mins). RT-LAMP was performed at 65°C for 30 mins, followed by measuring absorbance (OD) at 434 and 560 nm.

This difference between DNA and RNA could indicate the presence of RNAses or decomposition of the (chemically much less stable) RNA in the swab specimen due to heat incubation. However, IVT RNA that was added to heat incubated samples, followed by a second heat incubation, was still detected in the RT-LAMP assay (Figure 6d). Therefore, RNAses, and not heat, are the likely cause for the degradation of the IVT RNA. On the other hand, heat incubation is likely to inactivate RNAses, but does not affect the IVT RNA. Indeed, IVT RNA added after heat incubation could be detected in a subsequently conducted RT-LAMP assay with the RT-LAMP-specific sensitivity of approx 100 copies per µl sample (Figure 6e).

This analysis suggests that heat-inactivated swab specimens should be stable enough to allow detection of RNA, as long as no other components of these samples interfere with the RT-LAMP reaction.

In contrast to exogenous IVT RNA, the viral RNA in swab specimens of COVID-19 patients is present either inside cells or in viral capsids. This probably protects the RNA from RNAses until the sample is used for diagnostics. We next titrated a positive COVID-19 sample (CT=21) into control specimens from COVID-19 negative test subjects, alone or in the presence of 0.1% SDS. This SDS concentration was determined empirically to not disturb a subsequent RT-LAMP assay, when diluted 1:20 in the RT-LAMP mixture. The mixed extracts were then used for RT-LAMP, without (Figure 7a) and with (Figure 7b) heat treatment. In the absence of SDS, the viral RNA from the positive sample could still be detected at a dilution of 1:4096, while it was not detected at all in the presence of SDS, indicating that SDS leads to the release of RNAses and that the RNAs detected in the RT-LAMP assay are susceptible to degradation by these RNAses. We again tried to block degradation using an RNAse inhibitor that is active at 65°C (SUPERase-In RNase Inhibitor, Invitrogen), but without success (Figure 7).

**Figure 7:**
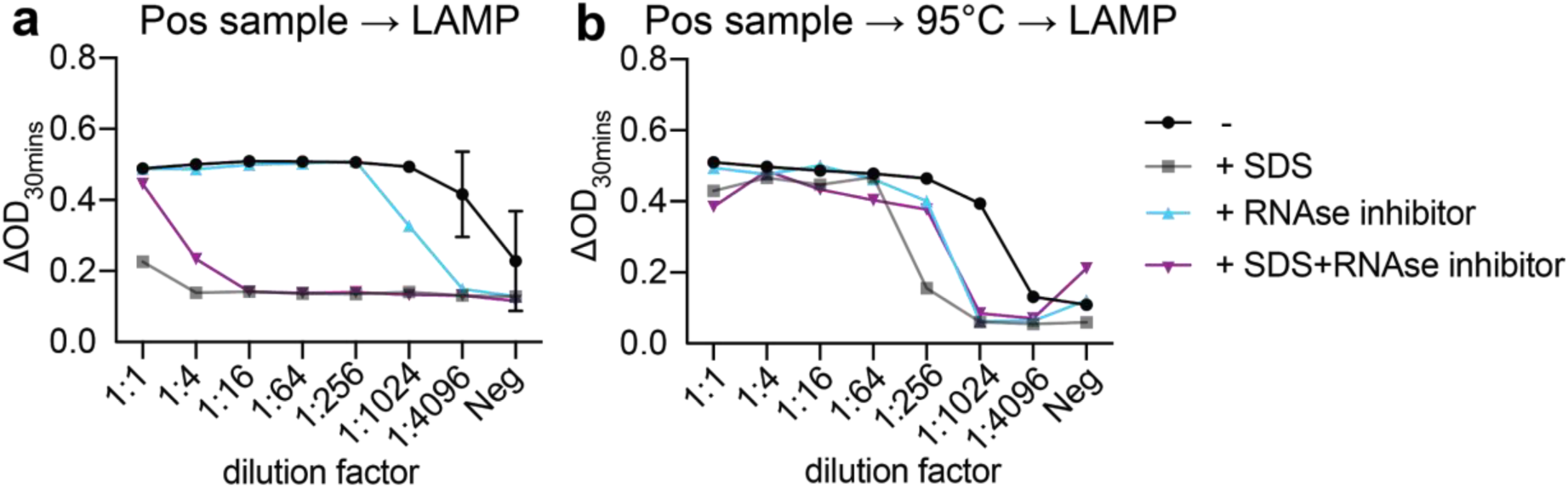
Swab-to-RT-LAMP assay titration of positive COVID-19 specimen. (**a**) A high titer positive COVID-19 specimen was diluted in control specimens and treated or not with 0.1% SDS and/or RNAse inhibitor as indicated, prior to RT-LAMP assay, n=2. (**b**) A high titer positive COVID-19 specimen was diluted in control specimens, heated (95°C / 5 min) and then treated or not as indicated, prior to RT-LAMP assay. RT-LAMP was performed at 65°C for 30 mins, followed by measuring absorbance (OD) at 434 nm and 560 nm.

These experiments established that, in addition to heat-treated swab specimens (hot swab-to-RT-LAMP assay), also ‘native’ swab specimens (direct swab-to-RT-LAMP assay) could be used directly for sensitive detection of viral RNA from a SARS-CoV-2 infection.

### Testing the swab-to-RT-LAMP assay on a collection of patient samples

Based on these preliminary experiments, we decided to use swab samples either directly, without any treatment, or after heat incubation for 5 min at 95°C. As an additional precaution we kept the samples in the cold (e.g. on ice) wherever possible. For testing large numbers of patient samples, we performed the assay in several 96-well plates. As shown in Figure 8 and Table 3, the majority of highly positive samples with CT < 30 induced a strong color change 30 mins into the RT-LAMP reaction. We have experienced that some positive samples do not induce a color change but did so when assayed a second time. We therefore recommend running this assay at least using technical duplicates.

**Figure 8:**
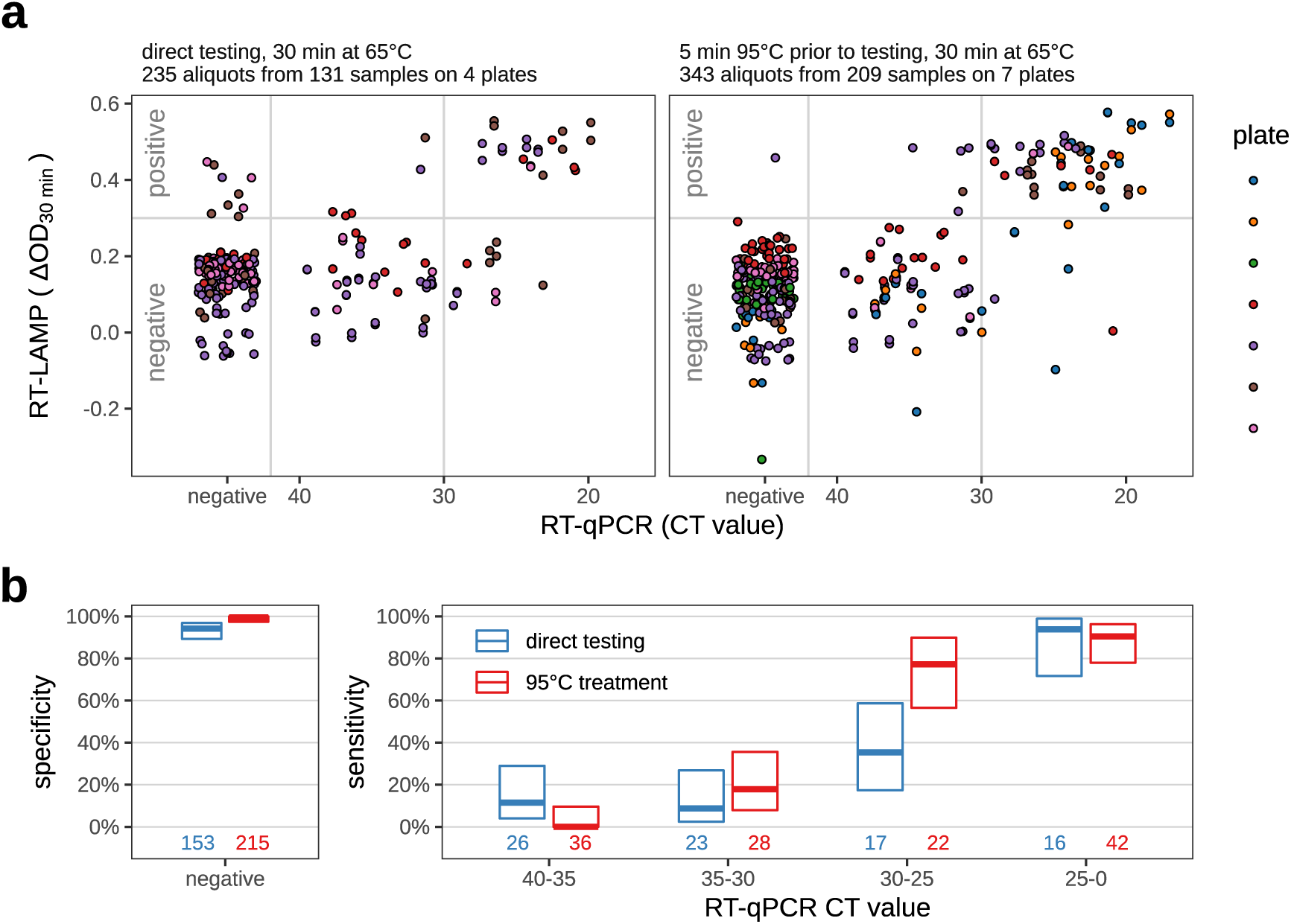
Direct swab-to-RT-LAMP assay on COVID-19 patient samples. (**a**) Scatter plot to visualize all samples of the swab-to-RT-LAMP assay test plates in one picture. Time point of measurement: 30 min after the start of the 65°C incubation. For the left panel, samples were assayed directly (direct swab-to-RT LAMP), for the right panel, the samples were first incubated at 95°C for 5 minutes, placed on ice and then used in the RT-LAMP assay (hot swab-to-RT LAMP). See Table 3 for counts. (**b**) Sensitivity and specificity of the RT-LAMP assay, as inferred from the data shown in (a) using the decision threshold indicated by the gray line in (a). Left: The specificity (i.e., the fraction of qPCR negatives correctly identified as negative by the RT-LAMP assay) and its 95% CI. Right: Thicker horizontal lines give, for several ranges of CT values, the sensitivity of the test (i.e., the proportion of samples within this CT range that the test detects as positive). The colored boxes indicate the 95% confidence intervals (Wilson’s binomial CI) for these values. Sample numbers are indicated at the bottom. For statistical parameters, see Supplementary Table S3.

For the direct swab-to-RT-LAMP assay overall specificity was 94.1 % (Wilson’s 95% CI: 89.2% - 96.9%), with a sensitivity for samples CT 0-25 93.8% (Wilson’s 95% CI: 71.7% - 98.9%) and for samples CT 25-30 35.3 % (Wilson’s 95% CI:17.3% - 58.7%).

For the hot swab-to-RT-LAMP, the overall specificity was 99.5 % (Wilson’s 95% CI: 97.4% - 99.9%) with a sensitivity for samples CT 0-25 90.5% (Wilson’s 95% CI: 77.9% - 94.9%) and for samples CT 25-30 77.3 % (Wilson’s 95% CI:56.6% - 77.8%). For more details, see Figure 8b and Supplementary Table S3. The heat treatment renders the assay more stringent with few false positives and at the same time more sensitive for samples CT 25-30 (defining a threshold at 0.3).

### Heterogeneity of pH specimen in swab-to-RT-LAMP assays

Comparison of the results of the swab-to-RT-LAMP assays with the isolated RNA revealed a much broader distribution of the ΔOD measurements in negative samples (Figure 9a, compared to Figure 3b). This is likely due to the patient-specific sample variability that influences the starting pH in the LAMP reaction. This might affect the interpretability of the measurement at 30 min (ΔOD_30min_). We investigated how this pH-shift influences the RT-LAMP assay. For three plates, the data acquired for the RT-LAMP assay also included measurements for time point 10 min (ΔOD_10min_). We plotted the change of the ΔOD between time points 10 min and 30 min (i.e. the difference ΔOD_30min_ - ΔOD_10min_, corresponding to the slope of the lines) versus ΔOD_30min_ (Figure 9b). This removed the variability of the values for samples that did not change the color (negative samples) and permitted a better separation of the positive from the negative samples.

**Figure 9:**
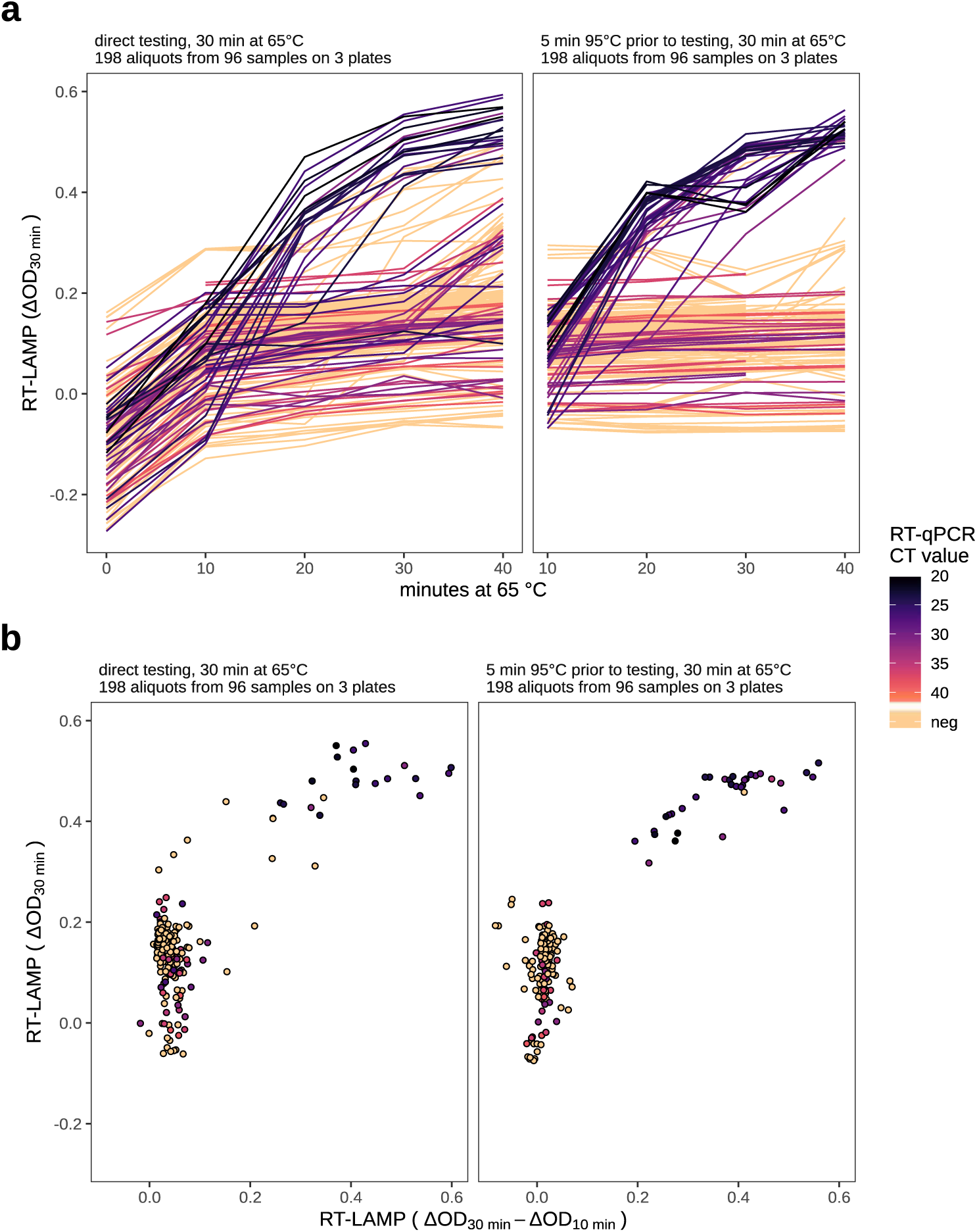
Scoring direct and hot swab-to-RT-LAMP by colorimetric changes over time. **(a)** Colorimetric read-out for swab-to-RT-LAMP, assessed every 10 minutes. Note the heterogeneity at the early time points. Heat treated samples are kept always on ice prior to incubation at 65°C. Therefore, we did not measure the ΔOD values at time point 0 min for this experiment. Also, time point 40 min was not available for one plate, and the kink in some lines of the right panel (at 30 min) was due to a transient equipment malfunction. **(b)** The samples shown here are those labelled in Figure 8 with the three colors at the bottom of the color legend.

We noticed that the pH-variability depended on the sample volume used for the RT-LAMP assay and the media composition of the swabs. For swabs in Amies medium (which was used for the clinical samples in this study) an RT-LAMP assay containing 1 µl of sample in a total volume of 20 µl was optimal.

In summary, the results obtained using native and heat-treated swab specimens from patients suggested better performance when using heat treatment of COVID-19 swab specimens prior to RT-LAMP assay.

## Discussion

### Primers

In this study we evaluated the use and suitability of RT-LAMP for the detection of SARS-CoV-2 infections in patients. We first compared primers and identified one primer set to detect SARS-CoV-2 N gene as the best, in agreement with Badhra et al., (2020) and Zhang et al., (2020). This coincides with the observation of Viehweger et al., (2020) that this gene showed the highest read coverage of all coronavirus genes when they sequenced RNA from tissue culture cells infected with coronavirus HCoV-229E. Using isolated IVT RNA, we established that the detection limit of RT-LAMP using N gene primers is approximately 100 copies, as reported by others (Lu et al., 2020).

### Decreased sensitivity of RT-LAMP in comparison to RT-qPCR

After establishing RT-LAMP for the detection of SARS-CoV-2 IVT RNA, we tested clinical samples. Using isolated RNA or direct swab specimens from COVID-19 patients, we found that RT-LAMP reliably detects SARS-CoV-2 IVT RNA in samples up to a CT ≈ 30 in RT-qPCR with a reported sensitivity and specificity values of RT-LAMP for SARS-CoV-2 detection in this study (99.7/ specificity, 92.0/ sensitivity for samples with CT < 30, Figure 4). Detection of SARS-CoV-2 RNA in samples from convalescent patients with low viral load and RT-qPCR test result with CT of > 30 often tested negative in the preceding or the subsequent RT-qPCR test. This could result from fluctuating viral load. During convalescence, Wölfel et al., (2020) detected less than one RNA molecule per µl of pharyngeal swab. Taken into account that the input of a typical RT-qPCR is equivalent to approximately 1-2% of a swab sample, this low number of molecules could result in a stochastic detection efficiency.

Based on our data we can conclude that RT-LAMP assays are suitable to recognize patients with a high viral load. On the other hand, for patients with low viral load (at the onset or later stages of disease), the sensitivity of RT-LAMP is insufficient to detect a SARS-CoV-2 infection. Whether alternative sampling, e.g. using sputum or stool, might be more reliable (Wölfel et al., 2020), is still under discussion, but it is certainly less practical for mass testing. Here, saliva samples collected according to the protocol of Wyllie et al., (2020) might offer advantages, if it turns out to be as sensitive as recent reports suggest.

### Direct and hot swab-to-RT-LAMP assays

While faster and more convenient, the direct swab-to-RT-LAMP assay is less sensitive and less robust than RT-LAMP using isolated RNA. To increase robustness, various treatments of crude swab samples are described (reviewed by Ribeiro da Silva et al., (2020)), many of which require additional processing of the samples by pipetting, as for example, adding Proteinase K to degrade contaminating proteins. However, Proteinase K has the same temperature optimum as the RT-LAMP reaction (65°C) and heat inactivation is not 100% efficient, even when incubating at 95°C for 10 min (see FAQ pages on www.qiagen.com), thus exposing the enzymes in the RT-LAMP reaction to unpredictable proteolytic influences. Rabe and Cepko (2020) suggest to use cheap silika preparation and novel sample inactivation protocols to enrich the RNA prior to LAMP, which however would come at the expense of complicating the simple swab-to-RT-LAMP workflow.

Finally, our analysis found that a short heat treatment of 5 min at 95°C, which poses minimal additional handling steps, does not destroy the RNA but rather stabilised it in the inactivated specimen, and improved the sensitivity and specificity of the swab-to-RT-LAMP assay (Figure 8). The heat likely helps to homogenize the sample, inactivate RNAses, and to break up the viral capsid to release the viral RNA.

Overall, our data clearly demonstrate the feasibility of swab-to-RT-LAMP testing and suggest applications especially in scenarios where no alternatives, such as RNA isolation before RT-LAMP or RT-qPCR tests, are available, e.g., due to shortages or in resource-poor settings. One promising lead for future applications is the exploration of hot swab-to-RT-LAMP with saliva specimens, which may show a higher sensitivity than in oropharyngeal swab specimens (To et al., 2020; Wyllie et al., 2020). Compatibility of RT-LAMP with direct saliva specimens has been shown before but only using spiked-in IVT RNA (Badhra et al., 2020, Rabe and Cepko 2020).

If RNA isolation is not an option, the hot swab-to-RT-LAMP performs best, whereas direct swab-to-RT-LAMP yields a substantial number of false positives, due to spurious amplification as also found elsewhere (Butler et al., (2020); Supplementary Figure S5, version 5 of the preprint). In general, while spike-in experiments with IVT RNA are somewhat informative, we have experienced clear differences when comparing to real swab specimens (Figure 6 and 7). We therefore strongly recommend validating any novel proposed rapid test for SARS-CoV-2 diagnostic using “real-life” clinical samples including a large fraction of negative clinical samples. To overcome the problem of spurious amplification, besides the solutions we propose here, also an expanded oligo set that incorporates sequence-specific probes (Bhadra et al., 2020) or a CRISPR/Cas12a based approach (Broughton et al., 2020) could be used. But these applications have yet to be tested with large numbers of diverse samples.

### Application of RT-LAMP for SARS-CoV-2 testing

With its good sensitivity for samples up to CT ≈ 30, the colorimetric RT-LAMP assay has several advantages to offer: It is fast, inexpensive, and it can be evaluated without any equipment. RT-LAMP reactions also appear to be less sensitive to contaminants in the sample; but care has to be taken that the used sample does not alter the pH, since the colorimetric RT-LAMP assay occurs under conditions of weak pH buffering. This is likely the bottleneck of the colorimetric RT-LAMP assay, which we found to work optimally with an input of 1 µl into the reaction. This corresponds to only 10% of the input used for the RT-qPCR. Some clinical samples contain high levels of contaminants which can lead to acidification of the reaction independent of the presence of a template RNA if too much sample is added (see also Supplementary Comments and Discussion for a detailed list of considerations and observations).

Diagnostic RT-qPCR tests usually include a technical internal control, i.e., another RNA species, which is spiked into all samples. This internal control is detected independent of the gene of interest to safeguard against the possibility of a general reaction failure within a sample tube. It would be desirable to have a similar precaution for RT-LAMP. Implementing such an approach for RT-LAMP with fluorescence read-out by multiplexing will likely be possible, but at the dispense of the simplicity of a colorimetric readout.

We see the potential role of RT-LAMP in the fight against the pandemic as follows: In settings where conventional diagnostic capacities are not exhausted, RT-qPCR tests with regulatory approval should be used for the diagnostic testing of suspected COVID-19 patients. Wherever available capacity allows so, regular testing of hospital personnel should also be continued with RT-qPCR, maybe in combination with sample pooling strategies to reach a higher throughput (Shani-Narkiss et al., 2020). Potential applications for RT-LAMP lie where the capacity of RT-qPCR cannot offer sufficient throughput as it is the case for needed mass screenings. Following the identification of one or few positive cases one can then apply “test, trace, and isolate” protocols using RT-qPCR to test this person’s contacts or everyone in their facility.

An important case here is sentinel testing: Especially in periods of the pandemic where social distancing has been relaxed due to a reduced rate of new infections, it is important to develop simple to implement instruments to test larger populations or selected cohorts. As we may expect that a local outbreak quickly comprises several individuals, it is more likely to detect at least one of them if priority is given to performing a large number of tests over using fewer but more sensitive tests.

### Application of LAMP-sequencing

We demonstrated a strategy to sequence the products of the LAMP reactions using multiplexed sample processing. Our particular implementation uses two sets of barcoded primers and is fully scalable so that in one sequencing run, many thousands of LAMP reactions can be quantitatively analyzed for the presence of viral genomic sequences. While we used Illumina dye sequencing on a NextSeq 550, more compartementalizable sequencing technologies, such as Oxford Nanopore Technologies sequencing, could be used for amplicon sequencing and counting (Buchmuller et al., 2019). The workflows shown here use LAMP-sequencing as a validation and backup procedure to double check the results of visual RT-LAMP assays. However, LAMP-sequencing could also facilitate upscaling of the workflows towards a direct analysis of many thousands of samples in an efficient manner, provided that an infrastructure is established that allows the collection of them. This could become an important part of workflows for routine testing of large populations.

The Zheng lab proposed decentralized LAMP assays using combinatorial primer barcoding and centralized mass analysis of LAMP products by next-generation sequencing as a means to upscale test-numbers dramatically (Schmid-Burgk et al., 2020). While this poses additional challenges in generating the LAMP reagents, it would reduce the challenge of sample handling on the analytical side.

In conclusion, we have comprehensively established how RT-LAMP could be used to implement testing for SARS-CoV-2 at a very high throughput, thus presenting a potentially highly effective instrument to combat the ongoing COVID-19 pandemic.

## Methods and materials

### Patient samples and sample handling

Specimens were collected as nasopharyngeal and oropharyngeal flocked swabs in Amies medium (eSwab, Copan Italia). Collected samples were transported in sterile containers, delivered to the diagnostic laboratory within a few hours, and then examined directly or stored at 4°C until further processing.

### RNA isolation and RT-qPCR

The standard diagnostic pipeline of the hospital laboratory was as follows: RNA was isolated from nasopharyngeal and oropharyngeal swab specimens using QIAGEN Kits (QIAGEN, Hilden, Germany); either automated on the QIASymphony (DSP Virus/Pathogen mini Kits) or QIAcube (QIAamp Viral RNA mini Kits) devices, or manually (QIAamp Viral RNA mini Kits), and eluted in 115 µl elution buffer. 10 µl of extracted sample was used in a 20 µl RT-qPCR reaction. RT-qPCR was carried out using various reagent mixes - LightMix Modular SARS and Wuhan CoV E-gene, LightMix Modular SARS and Wuhan CoV N-gene, LightMix Modular Wuhan CoV RdRP-gene and LightMix Modular EAV RNA Extraction Control (as internal Control) from TIB MOLBIOL Syntheselabor GmbH (Berlin, Germany) and LightCycler Multiplex RNA Virus Master (Roche, Germany) - according to manufacturer’s instructions. RT-qPCR was performed on a LightCycler 480 or 480 II (Roche, Germany). Thermal profile was as follows: reverse transcription step at 55°C for 5 min, followed by denaturation at 95°C for 5 min, and 45 amplification cycles (denaturation at 95°C 5 sec, annealing at 60°C 15 sec, and elongation at 72°C for 15 sec). For most incoming samples, PCR was performed first for gene E as a screen. Positive samples (CP≤ 40) were further analyzed for N gene and RdRP using the LightMix sets.

### RT-LAMP primer design and positive control

The RT-LAMP primer sets used in this study are designed against the ORF1a and N genes (Zhang et al., 2020) and were synthesized by Sigma-Aldrich (synthesis scale: 0.025 µmol, purification: desalt, solution: water). The sequences and the concentrations of each oligonucleotide in the 10x primer mix used for the RT-LAMP assay can be found in Supplementary Table S4.

An RNA positive control for N gene was obtained by amplifying a short fragment from 2019-nCoV_N_Positive Control plasmid (IDT, 10006625) with oligonucleotides T7-GeneN-Fragment.for and GeneN-Fragment.rev including the T7 promoter and a subsequent IVT with the MEGAscript T7 Kit (Invitrogen) purified using RNeasy MinElute Cleanup Kit (Qiagen).

### RT-LAMP assay

The assays were assembled in total reaction volumes of either 12.5 or 20 µl. Master mixes were prepared at room temperature for each reaction with 1x (6.25 or 10 µl respectively) of the WarmStart Colorimetric RT-LAMP 2X Master Mix (M1800, NEB) and 1x (1.25 or 2 µl respectively) of the 10x primer mix, filled up to 11.5 or 19 µl with nuclease-free water (AM9937, Ambion). 1 µl of sample was added and the reactions were incubated in a PCR cycler at 65°C for 15 - 60 min with the lid heated to 75°C. To take photographs or measurements at the indicated time points the reactions were taken out of the PCR cycler and placed into a 4°C cooling block for 30 sec to intensify the red and yellow color prior to the measurement. Photographs were taken with cell phone cameras.

### Quantification of the RT-LAMP reaction

Absorbance measurements of assays in 96-well PCR plates (4ti-0960/C, Brooks Life Sciences or 0030128672, Eppendorf) sealed with optically clear adhesive seals (GK480-OS, Kisker Biotech) were performed with a Spark Cyto or Infinite M200 (Tecan) at 434 and 560 nm with 25 flashes. These two peaks from phenol red are strongly changing during the acidification of the reaction (434 nm absorbance is increased, 560 nm absorbance is decreased), so we were subtracting the absorbance at 560 nm from the one at 434 nm to get a good readout of the color change.

### Swab-to-RT-LAMP assays

For direct and hot swab-to-RT-LAMP assays, patient swab specimens were transferred onto a 96-well plate. For direct swab-to-RT-LAMP assays, 1 µl of the patient specimens was added directly to the RT-LAMP mix in a ready-made plate with 19 µl LAMP mix per well. For hot swab-to-RT-LAMP assay, the plate with the specimens was sealed with a seal and heated in a PCR cycler for 5 mins at 95°C (with the lid heated to 105°C). The plate was cooled down to 4°C and 1 µl of the heat-treated patient specimens was quickly added to a ready-made plate with 19 µl LAMP mix per well. Plates with direct and hot swab-to-RT-LAMP assays were sealed, briefly spun down, and then incubated in a PCR cycler at 65°C for 10 - 60 min (with the lid heated to 75°C). To measure OD changes, the plates were removed at the indicated time points, quickly cooled down to 4°C, and measured using a Tecan reader (as described above). For time measurements, only the time at 65°C was counted.

### Data analysis

All data was analysed using R (R Core Team, 2020), using the tidyverse (Wickham et al., 2019), and ggplot2 system (Wickham, 2016), or using Graphpad Prism. Sensitivity and specificity values were obtained from count tables as indicated, binomial confidence intervals for these figures were calculated using Wilson’s method. The R code used to perform analyses and produce figures can be found on GitHub, together with well/sample-level data tables: https://github.com/anders-biostat/LAMP-Paper-Figures

### LAMP-sequencing procedure

Sequencing libraries for detecting viral sequences in RT-LAMP products were prepared by a modified Anchor-Seq protocol (Meurer et al., 2018; Buchmuller et al., 2019) using tagmentation instead of sonication for gDNA fragmentation (Picelli et al., 2014). The relevant primers are summarized in Supplementary Table S5.

In detail, transposon adapters containing well-defining barcodes and UMIs were annealed by mixing 25 µM of oligos (P5-UMI-xi501…5096-ME.fw, Tn5hY-Rd2-Wat-SC3) in 5 µM Tris-HCl (pH 8), incubating at 99°C for 5 min, and slowly cooling down to 20°C within 15 min in a thermocycler. Transposons were assembled by mixing 100 ng/µl Tn5(E54K, L372P) transposase (purified according to Hennig et al., 2018) with 1.25 µM annealed adapters in 50 mM Tris-HCl (pH 7.5) and incubating the reaction for 1 h at 23°C. Tagmentation was carried out by mixing 1.2 µl RT-LAMP product (~200 ng DNA) with freshly prepared tagmentation buffer (10 mM TAPS, pH 8.5; 5 mM MgCl_2_, 10 % (v/v) dimethylformamide) and 1.5 µl loaded transposase, and the reaction was incubated at 55°C for 10 min. Reactions were stopped by adding SDS to a final concentration of 0.033 %. Tagmented DNA of each plate was pooled and size-selected using a two-step AMPureXP bead (Beckman Coulter) purification to target for fragments between 300 and 600 bp. First, 50 µl pooled reaction was mixed with 50 µl water and bound to 55 µl beads to remove large fragments. To further remove small fragments, the supernatant of this reaction was added to 25 µl fresh beads and further purified using two washes with 80 % Ethanol before the samples were finally eluted in 10 µl 5 mM Tris-HCl, pH 8. One PCR per plate with 1 µl of the eluate and RT-LAMP-specific and Tn5-adapter-specific primers (P7nxt-GeneN-A-LBrc and P7-xi701..16, P5.fw) was performed using NEBNext Q5 HotStart polymerase (New England BioLabs) with two cycles at 62°C for annealing and 90 seconds elongation, followed by two cycles at 65°C for annealing and 90 seconds elongation, and 13 cycles at 72°C annealing and 90 seconds elongation. All PCR reactions were combined and 19 % of this pool was size-selected for 400-550 bp using a 2 % agarose/TAE gel and column purification (Macherey-Nagel). The final sequencing library was quantified by qPCR (NEB) and sequenced with a paired-end sequencing run on a NextSeq 550 machine (Illumina) with 20 % phiX spike-in and 136 cycles for the first read, 11 cycles to read the 11 nt long plate-index (i7) and 20 cycles to read the 11 nt-long well-index (i5) and the 9 nt-long UMI.

Reads were assigned to wells and counted using custom scripts. A read was considered as a match to SARS-CoV-2 N gene if at least one of three short sequences (~10 nt) not covered by RT-LAMP primers were observed, otherwise it was counted as unmatched. Sequencing reads were grouped by UMI and matched sequence positions with the aim to account for PCR amplification bias.

## Data Availability

data is available from https://github.com/anders-biostat/LAMP-Paper-Figures/

## Ethics statement

The intent of the work was for clinical methods development, as a response to the COVID-19 emergency. In this work we used pseudo-anonymized surplus material from samples that had been collected for clinical diagnostics of SARS-CoV-2. This is in accordance with the German Act concerning the Ethical Review of Research Involving Humans, which allows development and improvement of diagnostic assays using patient samples which were collected to perform the testing in question.

## Acknowledgments

We thank Vicent Pelechano (Karolinska), Xiushan Yin (Shenyang University of Chemical Technology), and Volker Lohmann (University of Heidelberg) for discussions and sharing unpublished work, Uta Merle for providing specimens and the Diagnostic department at University Hospital Heidelberg for providing isolated RNA from COVID-19 patients. We thank Vera Sonntag-Buck, Anke-Mareil Heuser, Ina Ambiel, Sophie Winter, Thorsten Müller, and Susanne Horner for excellent technical support. We thank Alexey Uvarovskii and Svetlana Ovchinnikova for IT support and Sylvia Kreger and Ursula Jäkle for additional technical help. VLDT and AF were supported by the Chica and Heinz Schaller foundation. DK was funded through internal funds from the DKFZ.

## Author contributions

Conceptualization: VLDT, MK, and SA, Methodology: VLDT, KH, MM, KB, DK, MK, and SA, Investigation: VLDT, KH, MM, KB, DK, AF, LPMK, DP, CG, SK, PC, DK, IBM, PS, MK, and SA, Formal Analysis: VLDT, KH, PC, LPMK, MK, and SA, Supervision: VLDT, KB, HGK, MK, and SA Administration: MK, HGK, IBM, KB, PS, and SA, Writing - Original Draft: VLDT, KH, MK and SA, Writing - Review and Editing: all authors.

## Supplementary Material

**Supplementary Table S1:**
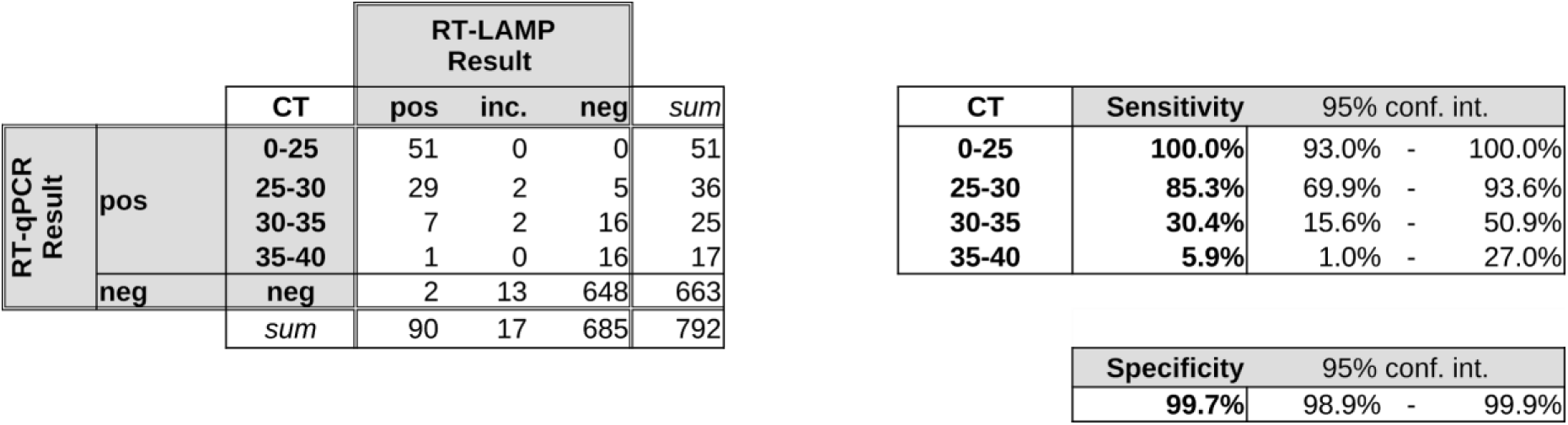
Cross tabulation of the results shown in Figure 4, comparing RT-qPCR and RT-LAMP test for 792 patient samples, stratified into CT value bins, and, inferred from this data, sensitivity (for each CT bin) and specificity of the LAMP test with respect to the qPCR results. “conf. int.” = Wilson’s 95% binomial confidence interval.

| RT-qPCR<br>Result |  | RT-LAMP<br>Result |  |  |  | sum |
| --- | --- | --- | --- | --- | --- | --- |
|  |  | CT | pos | inc. | neg |  |
|  | pos | 0-25 | 51 | 0 | 0 | 51 |
|  |  | 25-30 | 29 | 2 | 5 | 36 |
|  |  | 30-35 | 7 | 2 | 16 | 25 |
|  |  | 35-40 | 1 | 0 | 16 | 17 |
|  | neg | neg | 2 | 13 | 648 | 663 |
|  |  | sum | 90 | 17 | 685 | 792 |

| CT | Sensitivity | 95% conf. int. |  |
| --- | --- | --- | --- |
| 0-25 | 100.0% | 93.0% | 100.0% |
| 25-30 | 85.3% | 69.9% | 93.6% |
| 30-35 | 30.4% | 15.6% | 50.9% |
| 35-40 | 5.9% | 1.0% | 27.0% |

**Supplementary Table S2:**
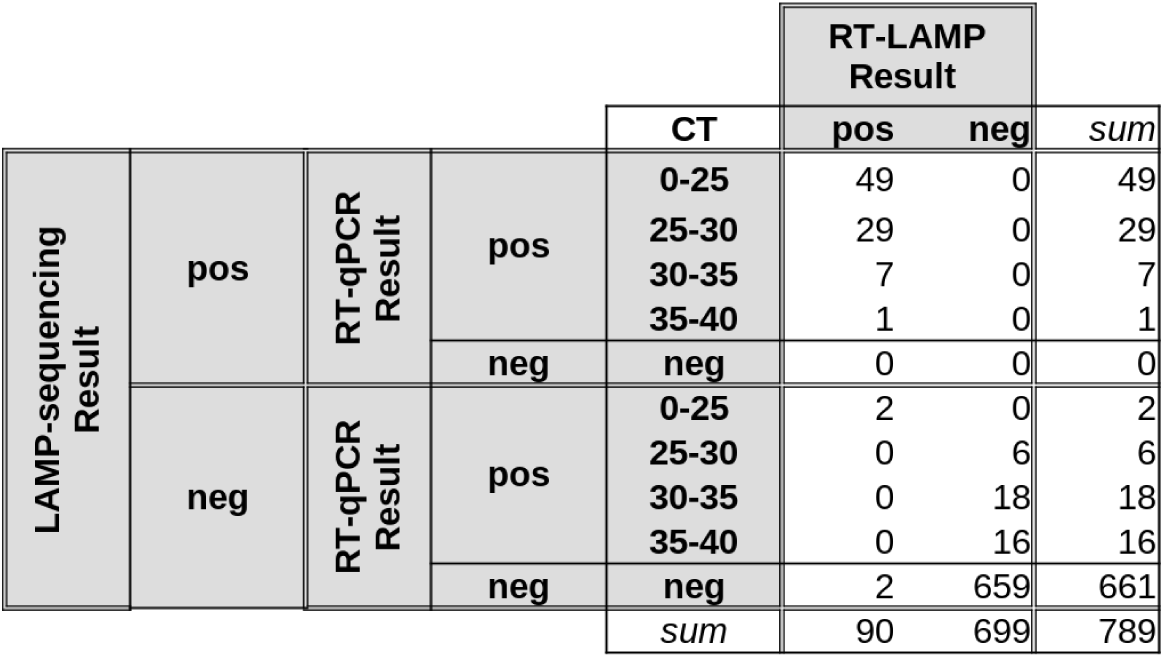
Cross-tabulation summarizing LAMP-sequencing results. (Figure 5): RT-qPCR and RT-LAMP results as in Supplementary Table 1. Numbers are split into cases where sequencing of RT-LAMP products showed virus sequences (LAMP-sequencing positive) and where they did not (LAMP-sequencing negative).

| LAMP-sequencing<br>Result |  |  |  | CT | RT-LAMP<br>Result |  | sum |
| --- | --- | --- | --- | --- | --- | --- | --- |
|  |  |  |  |  | pos | neg |  |
|  | pos | RT-qPCR<br>Result | pos | 0-25 | 49 | 0 | 49 |
|  |  |  |  | 25-30 | 29 | 0 | 29 |
|  |  |  |  | 30-35 | 7 | 0 | 7 |
|  |  |  |  | 35-40 | 1 | 0 | 1 |
|  |  |  | neg | neg | 0 | 0 | 0 |
|  | neg | RT-qPCR<br>Result | pos | 0-25 | 2 | 0 | 2 |
|  |  |  |  | 25-30 | 0 | 6 | 6 |
|  |  |  |  | 30-35 | 0 | 18 | 18 |
|  |  |  |  | 35-40 | 0 | 16 | 16 |
|  |  |  | neg | neg | 2 | 659 | 661 |
|  |  |  |  | sum | 90 | 699 | 789 |

**Supplementary Table S3:**
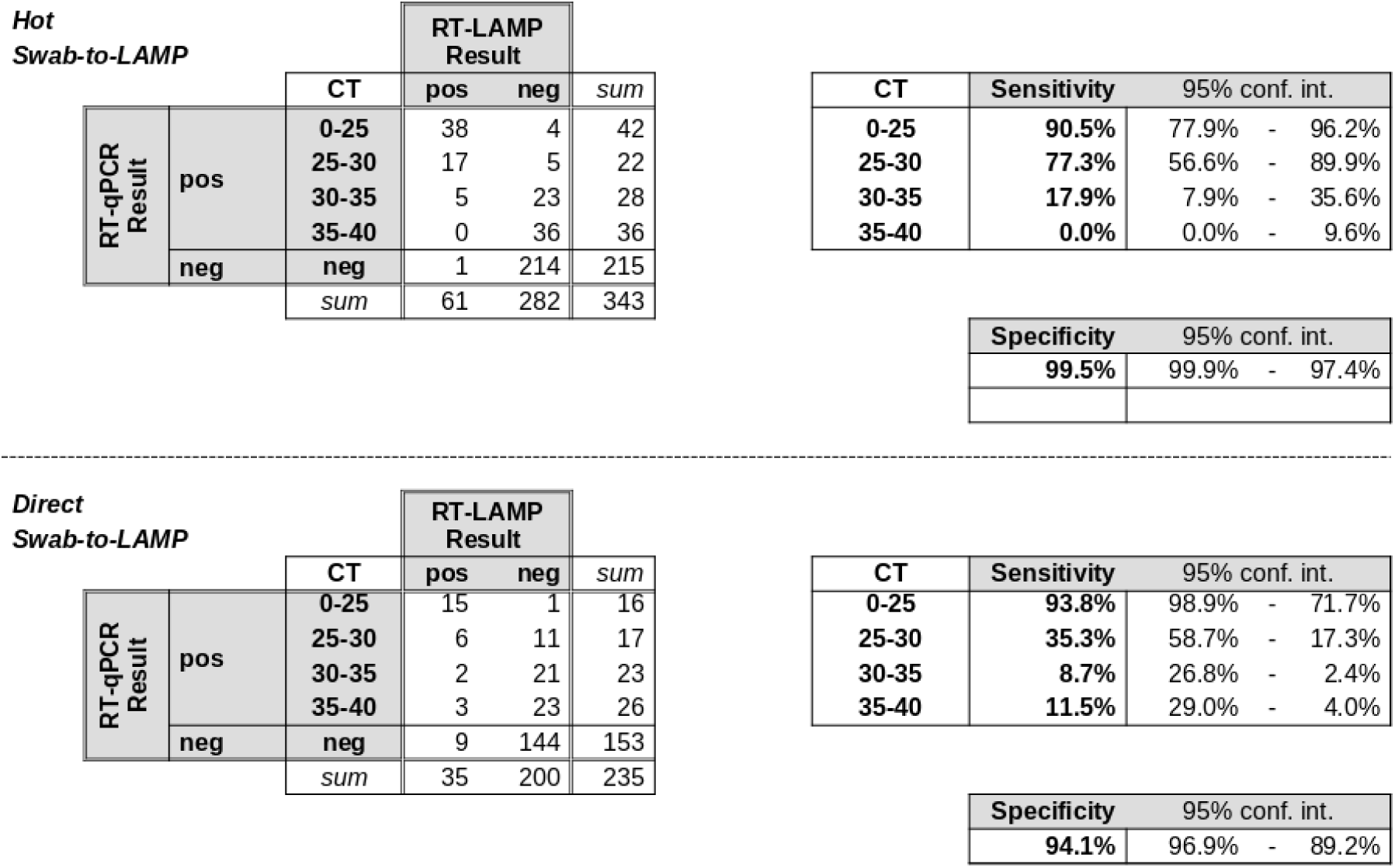
Cross tabulation of results from Figure 8 comparing RT-qPCR and RT-LAMP test for 592 patient samples, with sensitivity and specificity. “conf. int.” = Wilson’s 95% binomial confidence interval.

|  |  | CT | RT-LAMP Result |  | sum |
| --- | --- | --- | --- | --- | --- |
|  |  |  | pos | neg |  |
| RT-qPCR Result | pos | 0-25 | 38 | 4 | 42 |
|  |  | 25-30 | 17 | 5 | 22 |
|  |  | 30-35 | 5 | 23 | 28 |
|  |  | 35-40 | 0 | 36 | 36 |
|  | neg | neg | 1 | 214 | 215 |
|  |  | sum | 61 | 282 | 343 |

| CT | Sensitivity | 95% conf. int. |  |
| --- | --- | --- | --- |
| 0-25 | 90.5% | 77.9% | - 96.2% |
| 25-30 | 77.3% | 56.6% | - 89.9% |
| 30-35 | 17.9% | 7.9% | - 35.6% |
| 35-40 | 0.0% | 0.0% | - 9.6% |

| Specificity | 95% conf. int. |  |
| --- | --- | --- |
| 99.5% | 99.9% | - 97.4% |

Direct Swab-to-LAMP
|  |  | CT | RT-LAMP Result |  | sum |
| --- | --- | --- | --- | --- | --- |
|  |  |  | pos | neg |  |
| RT-qPCR Result | pos | 0-25 | 15 | 1 | 16 |
|  |  | 25-30 | 6 | 11 | 17 |
|  |  | 30-35 | 2 | 21 | 23 |
|  |  | 35-40 | 3 | 23 | 26 |
|  | neg | neg | 9 | 144 | 153 |
|  |  | sum | 35 | 200 | 235 |

| CT | Sensitivity | 95% conf. int. |  |
| --- | --- | --- | --- |
| 0-25 | 93.8% | 98.9% | - 71.7% |
| 25-30 | 35.3% | 58.7% | - 17.3% |
| 30-35 | 8.7% | 26.8% | - 2.4% |
| 35-40 | 11.5% | 29.0% | - 4.0% |

**Supplementary Table S4:**
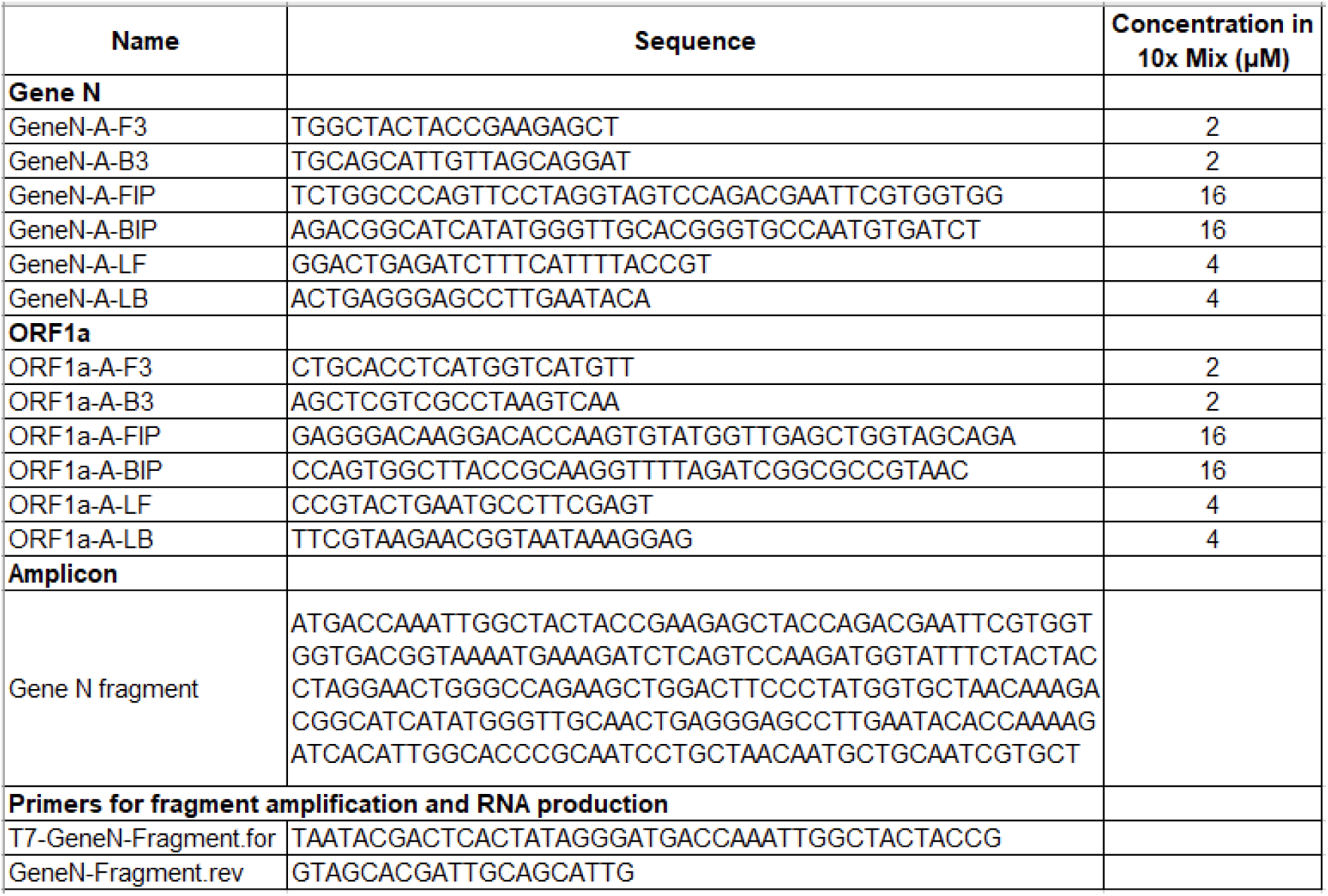
Sequences of amplicons and RT-LAMP primer sets

**Supplementary Table S5:** Sequences of primers used for LAMP-sequencing (provided as separate data file).

**Supplementary Figure S1:**
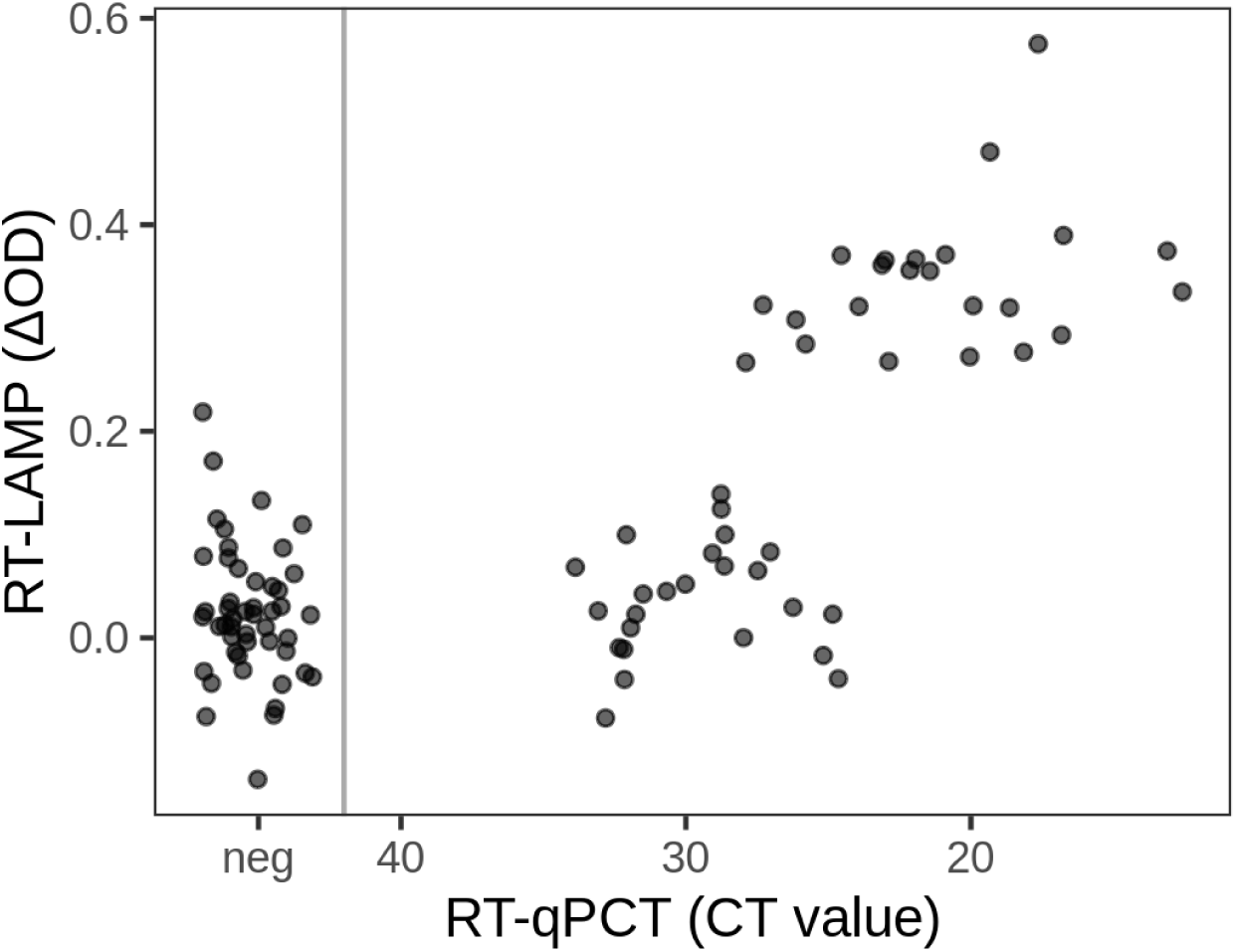
Comparison of the RT-LAMP results with the CT values from RT-qPCR. **(d)** Same samples as in (Figure 3c) using a primer set for the 1a gene for time point 40 minutes.

**Supplementary Figure S2:**
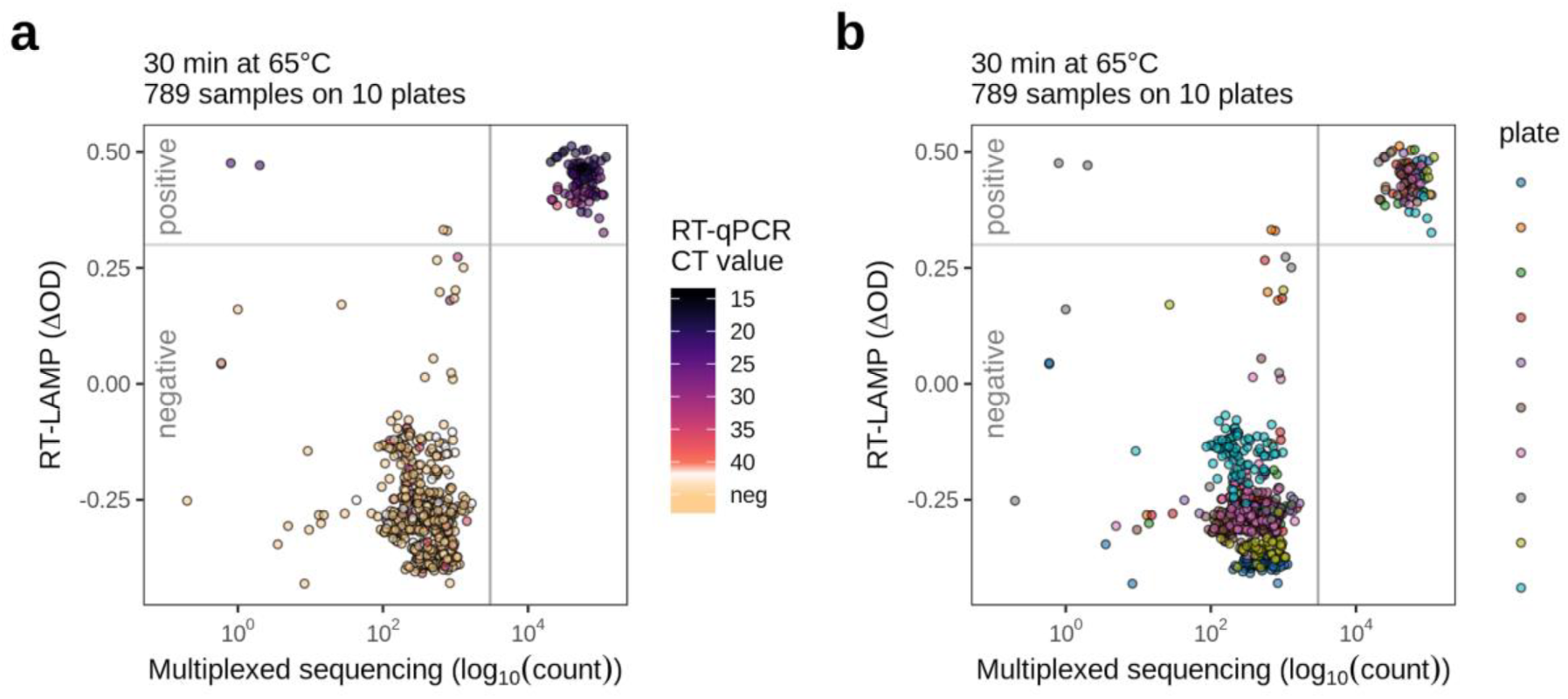
Multiplexed sequencing count data used for Figure 5 stratified by RT-qPCR-derived CT values **(a)** or processed sample plate **(b)**. RT-LAMP assay values were read after incubating 30 min at 65°C. A sequencing count of 3000 virus-matching sequences (grey vertical line) was used to flag a sample as virus-positive.

## Supplementary Discussion and Comments

### Important considerations when using RT-LAMP assays and reagents

1. The buffer capacity of the WarmStart Colorimetric RT-LAMP 2X Master Mix (DNA & RNA) from NEB (M1800) is very low in order to allow a color change as a function of synthesized DNA. High sample volumina, buffered samples and compounds influencing the buffer capacity of the reaction can yield false positive or false negative results.
2. The LAMP reaction can yield non-specific products which induce a pH shift; these appear with a delay of approx 10 min and thus measurements at time points > 35 min are prone to contain increasing numbers of false positives.
3. Best results were obtained with freshly prepared RT-LAMP master mixes that were kept on ice in a closed tube prior to the addition of the sample
4. Colorimetric RT-LAMP mastermixes are not very stable. Old master mixes (set up with primers) are characterized by a color change in the negative control sample already at the time point 30 min, or even earlier. This yields false positive results. Freshly prepared mastermixes can be stored maximally 14-18 hours at −20°C. Longer storage also yields false positive results.
5. Exposure to the atmosphere (open lid) leads to a slow drop in pH, presumably due to acidification by atmospheric CO_2_. Pipetting of RT-LAMP mixes into 96 well plates needs to be done immediately before use. Keep plates on ice while adding the samples. Pipette carefully and avoid bubbles which, if they burst, could contaminate adjacent samples.

### Sample and reaction volume

6. For purified RNA samples we use 1 µl of RNA in a total reaction volume of 12.5 µl. Larger quantities of the RNA (e.g. 4 µl) did not significantly improve detection sensitivity (probably due to impurities in the RNA preparations, e.g. EtOH).
7. For swab-to-RT-LAMP assays we use 1 µl of sample in a total reaction volume of 20 µl. Likewise, larger quantities of sample input did not significantly improve detection sensitivity, likely due to the presence of undefined inhibitors.

### Stability of color change (see also points 1-4)

8. Swab samples in Amies medium are slightly acidic and, depending on the patient, we observed a drop in pH in the reaction mixture already before the reaction started (see also Figure 9). Using 0.9% saline solution (instead of the Amies medium used in clinical samples) does not lead to a pH drop at the time point 0 min.

### Quantification of the color change

9. The RT-LAMP test, like the qPCR, is based on a time-dependent reaction. Our experiments show that the measurement is best between 27-30 min and we propose to implement this time point for read-out.
10. Documentation of the color change can be done in many ways. While we used a Tecan plate scanner for OD measurements at two wavelengths, we found that placing the test strips or plates on a color scanner or copy machine with scanner function is another very reliable and reproducible way to document results. Hue measurements can then be used to quantify the results. Cameras of mobile phones work also well (Figure 2a), but reflection of light and perspective distortions make such images less reproducible and difficult to quantify.

### Contaminations

11. When pipetting PCR strips, always close the lids to avoid contaminations. When using 96 well plates, pipette carefully and absolutely avoid bubbles which, if they burst, could contaminate adjacent samples.
12. Successful RT-LAMP reactions contain large quantities of DNA that are a perfect template for the assay. Therefore, after the reaction the tubes should not be opened and be carefully disposed of.
13. Analysis of RT-LAMP reactions for gel electrophoresis requires that the tubes are opened. This should be done in a well separated location and using equipment that is not shared with work benches where RT-LAMP reactions are set up or where patient samples are manipulated.

